# High-dimensional mass cytometry analysis of NK cell alterations in Acute Myeloid Leukemia identifies a subgroup with adverse clinical outcome

**DOI:** 10.1101/2020.10.01.20204867

**Authors:** Anne-Sophie Chretien, Raynier Devillier, Samuel Granjeaud, Charlotte Cordier, Clemence Demerle, Nassim Salem, Julia Wlosik, Florence Orlanducci, Emilie Gregori, Magali Paul, Philippe Rochigneux, Thomas Pagliardini, Mathieu Morey, Cyril Fauriat, Nicolas Dulphy, Antoine Toubert, Herve Luche, Marie Malissen, Didier Blaise, Jacques A. Nunès, Norbert Vey, Daniel Olive

**Affiliations:** Team Immunity and Cancer, Centre de Recherche en Cancérologie de Marseille (CRCM), Inserm, U1068, CNRS, UMR7258, Institut Paoli-Calmettes, Aix-Marseille University, UM105, Marseille, France; Immunomonitoring department, Institut Paoli-Calmettes, Marseille, France; Hematology Department, Centre de Recherche en Cancérologie de Marseille (CRCM), Inserm, U1068, CNRS, UMR7258, Institut Paoli-Calmettes, Aix-Marseille University, UM 105, Marseille, France; Systems Biology Platform, Centre de Recherche en Cancérologie de Marseille (CRCM), Inserm, U1068, CNRS, UMR7258, Institut Paoli-Calmettes, Aix-Marseille University, UM 105, Marseille, France; Biopathology Department, Institut Paoli-Calmettes, Marseille, France; Centre d’Immunophénomique–Luminy (Ciphe), Inserm US012, CNRS UMS3367, Aix-Marseille University, Marseille, France; ImCheck therapeutics, Marseille, France; Medical Oncology Department, Institut Paoli-Calmettes, Marseille, France; Datactivist, Aix-en-Provence, France; Université Paris Diderot, Sorbonne Paris Cité, Institut Universitaire d’Hématologie, Immunology and Histocompatibility department, Hôpital Saint-Louis, APHP, Inserm UMRS-1160, Paris, France; Centre d’Immunologie de Marseille-Luminy (CIML), Inserm CNRS, Aix-Marseille University, Marseille, France

**Keywords:** AML, natural killer cells, CD56^-^CD16^+^ NK cells, prognostic biomarkers, mass cytometry

## Abstract

Natural killer (NK) cells are major anti-leukemic immune effectors. Leukemic blasts have a negative impact on NK cell function and promote the emergence of phenotypically and functionally impaired NK cells. In the present work, we highlight an accumulation of CD56^-^CD16^+^ unconventional NK cells in acute myeloid leukemia (AML), an aberrant subset initially described as being elevated in patients chronically infected with HIV-1. Deep phenotyping of NK cells was performed using peripheral blood from patients with newly-diagnosed AML (N=48, HEMATOBIO cohort, NCT02320656) and healthy subjects (N=18) by mass cytometry. We evidenced a moderate to drastic accumulation of CD56^-^ CD16^+^ unconventional NK cells in 27% of patients. These NK cells displayed decreased expression of NKG2A as well as the triggering receptors NKp30, and NKp46, in line with previous observations in HIV-infected patients. High-dimensional characterization of these NK cells highlighted a decreased expression of three additional major triggering receptors required for NK cell activation, NKG2D, DNAM-1, and CD96. A high proportion of CD56^-^CD16^+^ NK cells at diagnosis was associated with an adverse clinical outcome, with decreased overall survival (HR=0.13; P=.0002) and event-free survival (HR=0.33; P=.018), and retained statistical significance in multivariate analysis. Pseudo-time analysis of the NK cell compartment highlighted a disruption of the maturation process, with a bifurcation from conventional NK cells toward CD56^-^CD16^+^ NK cells. Overall, our data suggest that the accumulation of CD56^-^CD16^+^ NK cells may be the consequence of immune escape from innate immunity during AML progression.

**Significance:** This work provides the first report of accumulation of unconventional CD56-CD16+ NK cells in non-virally induced malignancies. Pseudotime analysis highlights a bifurcation point occurring during the course of NK cell maturation, providing elements regarding the possible origin of CD56-CD16+ NK cells. Increased frequency of CD56-CD16+ NK cells is associated with adverse clinical outcome in AML and might contribute, as well as other maturation defects, to a defective control of AML progression. Overall, accumulation of CD56-CD16+ NK cells could be an important feature of immune escape from innate immunity.

**Graphical abstract:** 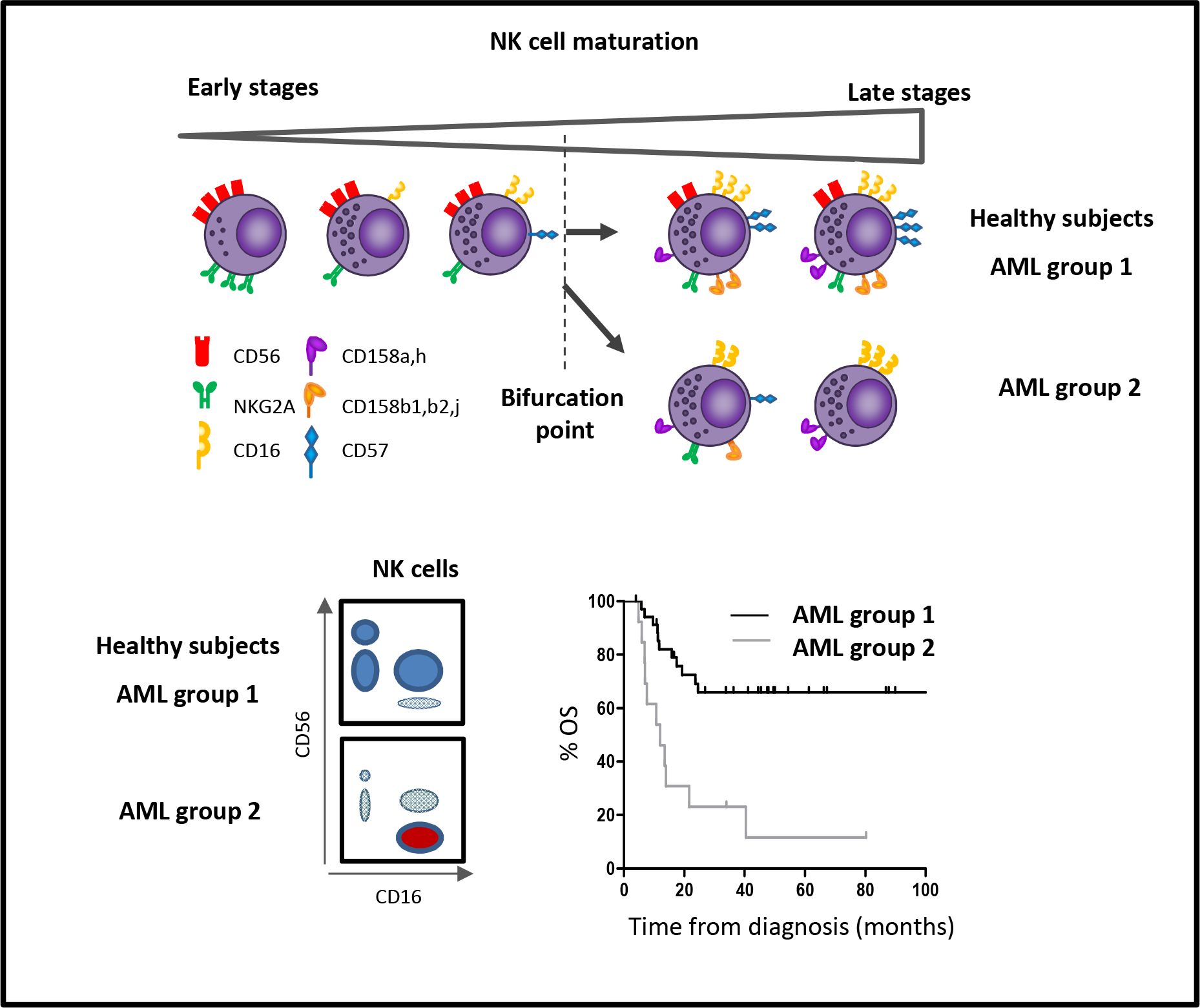

*Key points:*

- A disruption in the maturation process of NK cells leads to accumulation of unconventional CD56^-^ CD16^+^ NK cells in patients with AML
- High frequency of CD56^-^CD16^+^ NK cells is associated with adverse clinical outcome

## INTRODUCTION

Natural killer (NK) cells are critical cytotoxic effectors involved in leukemic blast recognition, tumor cell clearance, and maintenance of long-term remission.^1^ NK cells directly kill target cells without prior sensitization, enabling lysis of cells stressed by viral infections or tumor transformation. NK cells are divided into different functional subsets according to CD56 and CD16 expression.^2–4^ CD56^bright^ NK cells are the most immature NK cells found in peripheral blood. This subset is less cytotoxic than mature NK cells and secretes high amounts of chemokines and cytokines such as IFNγ and TNFα. These cytokines have a major effect on the infected or tumor target cells, and play a critical role in orchestration of the adaptive immune response through dendritic cell activation. CD56^dim^CD16^+^ NK cells, which account for the majority of circulating human NK cells, are the most cytotoxic NK cells. NK cell activation is finely tuned by integration of signals from inhibitory and triggering receptors, in particular NKp30, NKp46, and NKp44, DNAM-1, and NKG2D.^5^ Upon target recognition, CD56^dim^CD16^+^ NK cells release perforin and granzyme granules and mediate antibody-dependent cellular cytotoxicity through CD16 (FcγIIIR) to clear transformed cells.

NK cells are a major component of the anti-leukemic immune response, and NK cell alterations have been associated with adverse clinical outcomes in acute myeloid leukemia (AML).^6–9^ Therefore, it is crucial to better characterize AML-induced NK cell alterations in order to optimize NK cell-targeted therapies. During AML progression, NK cell functions are deeply altered, with decreased expression of NK cell-triggering receptors and reduced cytotoxic functions, as well as impaired NK cell maturation.^6,9–13^ Cancer-induced NK cell impairment occurs through various mechanisms of immune escape, including shedding and release of ligands for NK cell-triggering receptors, release of immunosuppressive soluble factors such as TGFβ, adenosine, PGE2, or L-kynurenine, and interference with NK cell development, among others.^14^

Interestingly, these mechanisms of immune evasion are also seen to some extent in chronic viral infections, notably HIV.^2^ In patients with HIV, NK cell functional anergy is mediated by the release of inflammatory cytokines and TGFβ, the presence of MHC^low^ target cells, and the shedding of ligands for NK cell-triggering receptors.^2^ As a consequence, some phenotypical alterations described in cancer patients are also induced by chronic HIV infections, with decreased expression of major triggering receptors such as NKp30, NKp46, and NKp44,^15,16^ decreased expression of CD16,^17^ and increased expression of inhibitory receptors such as TIGIT^18^ all observed. In addition, patients with HIV display an accumulation of CD56^-^CD16^+^ unconventional NK cells, a highly dysfunctional NK cell subset.^19,20^ Mechanisms leading to the loss of CD56 are still poorly described, and the origin of this subset of CD56^-^ NK cells is still unknown. To date, two hypotheses have been considered: CD56^-^ NK cells could be terminally differentiated cells arising from a mixed population of mature NK cells with altered characteristics, or could expand from a pool of immature precursor NK cells.^21^ Expansion of CD56^-^CD16^+^ NK cells is mainly observed in viral non-controllers.^19,20^ Indeed, CD56 is an important adhesion molecule involved in NK cell development, motility and pathogen recognition.^22–27^ CD56 is also required for the formation of the immunological synapse between NK cells and target cells, lytic functions, and cytokine production.^26,28^ As a consequence, CD56^-^CD16^+^ NK cells display lower degranulation capacities, decreased expression of triggering receptors, perforin, and granzyme B, dramatically reducing their cytotoxic potential, notably against tumor target cells.^2,19,20,29,30^ In line with this loss of the cytotoxic functions against tumor cells, patients with concomitant Burkitt lymphoma and Epstein Barr virus infection display a dramatic increase of CD56^-^CD16^+^ NK cells,^30^ which could represent an important hallmark of escape to NK cell immunosurveillance in virus-driven hemopathies.

To our knowledge, this population has not been characterized in the context of non-virally induced hematological malignancies. In the present work, we investigated the presence of this population of unconventional NK cells in patients with AML, its phenotypical characteristics, and the consequences of its accumulation on disease control. Finally, we explored NK cell developmental trajectories leading to the emergence of this phenotype.

## PATIENTS AND METHODS

### Patients and study design

This monocentric study (Paoli-Calmettes Institute, Marseille, France) included forty-eight patients with newly diagnosed non-acute promyelocytic leukemia AML from the HEMATOBIO cohort (NCT02320656). Patients were aged 19 to 80 years (mean ±SD = 54.0±14.0). All patients received cytarabine and anthracycline–based induction chemotherapy as previously described.^43^ Eight patients (16.7%) were treated with allogeneic stem cell transplantation as a consolidation therapy. Patients’ characteristics are summarized in Table 1.

### Ethics statement

All participants gave written informed consent in accordance with the Declaration of Helsinki. The entire research procedure was approved by the institutional review board of the Paoli-Calmettes Institute.

### Clinical samples

Peripheral blood mononuclear cells (PBMCs) cryopreserved in 90% albumin/10% DMSO were obtained before (N=48) and after induction chemotherapy (N=16, paired samples), and from age-matched healthy volunteers (N=18). Handling, conditioning, and storage of samples were performed by the Paoli-Calmettes Tumor bank, which operates under authorization # AC-2007-33 granted by the French Ministry of Research.

### Mass cytometry analysis

PBMCs were thawed and processed as previously described.^6^ PBMCs were washed with RPMI 1640 medium with 10% fetal calf serum and incubated in RPMI 1640 with 2% fetal calf serum and 1/10000 Pierce^®^ Universal Nuclease 5kU (Thermo Fisher Scientific, Waltham, MA, USA) at 37°C with 5% CO_2_ for 30 min. Cells were centrifuged and incubated with cisplatin 0.1 M to stain dead cells. Aspecific epitopes were blocked with 0.5 mg/mL Human Fc Block (BD Biosciences, San Jose, CA, USA). Two million PBMCs were stained for 45 min at 4°C with the extracellular antibodies (Supplemental Table 1). Cells were centrifuged and barcoded with the Cell-ID™ 20-Plex Pd *Barcoding Kit* (Fluidigm, San Francisco, CA, USA) according to the manufacturer’s recommendations. Cells were washed and samples were combined and stained with metal-labeled anti-phycoerythrin secondary antibodies for 30 min at 4°C. After centrifugation, cells were washed and permeabilized with the Foxp3 Staining Buffer Set (eBioscience, San Diego, CA, USA) for 40 min at 4°C. Intracellular aspecific epitopes were blocked with 0.5 mg/mL Human Fc Block for 40 min at 4°C before incubation with the mix of intracellular antibodies for 40 min at 4°C in Foxp3 Staining Buffer (Supplemental Table 1). Cells were then washed and labeled overnight with 125 nM iridium intercalator (Fluidigm) in Cytofix (BD Biosciences). Finally, cells were diluted in EQTM Four Element Calibration Beads (Fluidigm) before acquisition on a Helios^®^ instrument (Fluidigm).

### Statistical analysis

Statistical analyses were carried out using Graph Pad Prism V5.01 (San Diego, CA, USA) and SPSS V9.0 (Chicago, IL, USA). The χ^2^ or Fisher’s exact test was used to assess association between variables. For multiple comparisons of paired values, a Friedman test was performed followed by a Dunn’s post-test. Groups of patients with a high or low frequency of CD56^-^CD16^+^ NK cells were defined based on optimized cut-points using maximally selected log-rank statistics (maxstat package, R software V 3.6.2)^44^ and on imposing that no group could represent fewer than 25% of patients. For survival analyses, overall survival (OS) was defined as the time from diagnosis until death from any cause, and event-free survival (EFS) as the time between induction and relapse, death from any cause, or induction failure, whatever occurred first. Patients without an event were censored at the time of their last follow-up. Survival times were estimated by the Kaplan–Meier method and compared using the log-rank test. A multivariate Cox regression model was used to assess the prognostic value of CD56^-^CD16^+^ NK cells while adjusting for other prognostic factors. Candidate variables for the Cox regression were European Leukemia Net genetic classification, age at diagnosis, leukocytosis, and percentage of CD56^-^CD16^+^ NK cells. Continuous variables were discretized as follows: age < or ≥50 years old; leukocytosis: < or ≥50 G/L; CD56^-^CD16^+^ NK cells: < or ≥10%. All factors with a *P* value <.15 in univariate analysis were considered to be candidates for the backward stepwise Cox regression model. For subgroup analyses, patients were divided into two groups, a relapsed AML group and a long-term complete remission group, according to clinical outcome after 24 months of follow-up. The limit of significance was set at P<.05.

### Algorithm-based high-dimensional analysis

NK cells were manually defined as CD13^-^CD33^-^CD34^-^CD45^+^CD3^-^CD19^-^CD56^+^ or CD13^-^CD33^-^CD34^-^CD45^+^ CD3^-^CD19^-^CD56^-^CD16^+^ and exported using FlowJo V10.6.2. The gating strategy is displayed in Supplemental Figure 1. Data were arcsinh-transformed with a cofactor of 5. NK cell populations were automatically defined using hierarchical stochastic neighbor embedding (h-SNE) analysis (Cytosplore V2.2.1) with default settings (30 perplexity and 1,000 iterations).^45^ For the h-SNE analyses, consensus files were generated for each group of patients with a fixed number of NK cells in order to obtain a representative and balanced view of all patient groups. NK cell trajectory inferences were performed using the Wishbone algorithm.^32^ The library was updated for Python V3.6 and is available at https://github.com/moreymat/wishbone/releases/tag/0.4.2-py.3.

## RESULTS

### Accumulation of CD56^-^CD16^+^ NK cells in patients with AML

PBMCs from 48 newly diagnosed AML patients were analyzed by mass cytometry. Based on expression of CD56 and CD16, we defined four NK cell subsets: CD56^bright^ NK cells, CD56^dim^CD16^-^ NK cells, CD56^dim^CD16^+^ NK cells, and CD56^-^CD16^+^ NK cells (Supplemental Figure 1). In 5 out of 48 patients (10.4%), we observed a massive accumulation of CD56^-^CD16^+^ NK cells, with more than 20% of total NK cells represented in this cluster. In 8 out of 48 patients (16.7%), we observed a moderate accumulation of CD56^-^CD16^+^ NK cells, with 10–19% of total NK cells represented in this cluster (Figure 1A). The threshold of 10% was defined on the basis of the maxstat algorithm (Supplemental Figure 2). Patients with ≤10% CD56^-^CD16^+^ NK cells are referred to as group 1 (N=35) and patients with >10% CD56^-^CD16^+^ NK cells are referred to as group 2 (N=13) in the rest of the article (Figure 1A). The frequency of CD56^-^CD16^+^ NK cells was significantly higher in group 2 than in group 1 or in healthy volunteers, with mean frequencies of 24.9% *vs* 3.5% and 7.7%, P<.001 and P<.05, respectively (Figure 1B). This threshold is illustrated on the intensity distribution graph using a kernel density estimation (Figure 1C).

**Figure 1.**
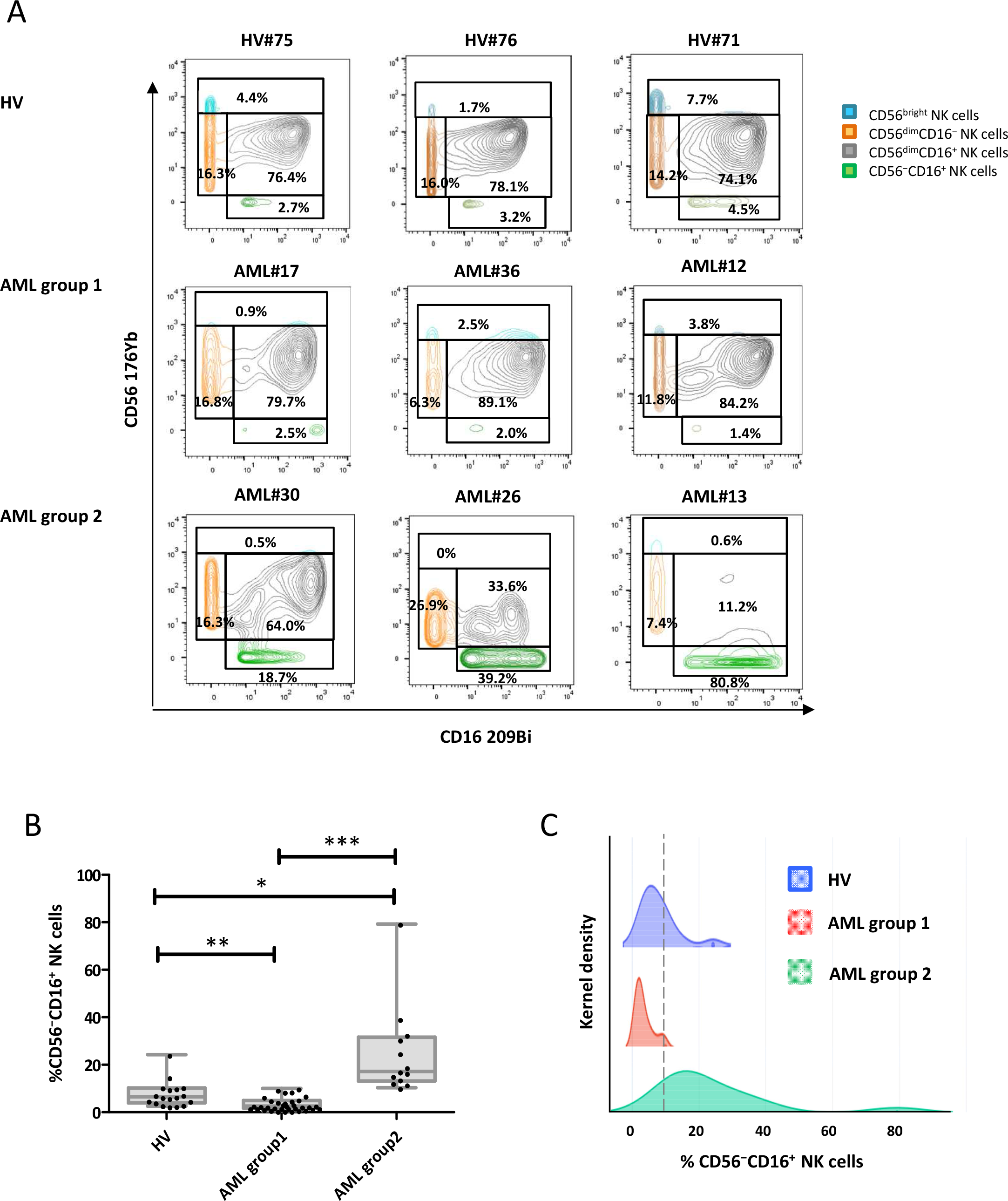
Accumulation of unconventional CD56^-^CD16^+^ NK cells in AML. PBMC from 48 newly-diagnosed AML patients and 18 HV were phenotyped by mass cytometry. (A) NK cell phenotype by CD56 and CD16 expression; representative examples of HV and AML patients without (group 1) or with (group 2) accumulation of CD56^-^CD16^+^ NK cells. (B) Frequency of CD56^-^CD16^+^ NK cells in HV and patients with AML. Results are presented as interquatile ranges, median, and whiskers from minimum to maximum. (C) Threshold visualization and value distribution illustrated by Kernel density estimation. HV, healthy volunteer. *: P<0.05; **: P<0.01; ***: P<0.001.

More than 95% of cells expressed the transcription factors T-bet and Eomes across the four NK cell subsets (Supplemental Figure 3A). However, as group 1 innate lymphoid cells (ILC1) share expression of T-bet with NK cells,^31^ we assessed expression of the ILC markers CD25, CD127 (IL7R), and CD278 (ICOS) to exclude the hypothesis that ILC1s were accumulated in patients in group 2. As expected, CD25 and IL7R expression was mostly restricted to CD56^bright^ NK cells, whereas expression was reduced in CD56^dim^ and CD56^-^ NK cells compared with other clusters of conventional CD56^+^ NK cells (Supplemental Figure 2B). In addition, more than 95% of NK cells were ICOS^-^, including those within the CD56^-^CD16^+^ subset, further confirming that CD56^-^CD16^+^ cells were not ILC1s (Supplemental Figure 3B).

### High-dimensional analysis of CD56^-^CD16^+^ NK cells phenotypical characteristics

Loss of CD56 expression on NK cells has been associated with a loss of two of the main NK cell-triggering receptors, NKp30, NKp46, as well as NKG2A in HIV-infected patients.^20^ We used the h-SNE algorithm to explore NK cell alterations associated with accumulation of CD56^-^CD16^+^ NK cells in AML. As described in HIV infection, CD56^-^CD16^+^ NK cells from patients with AML also exhibited a loss of NKp30, NKp46, and NKG2A (Figure 2A and B). In addition, h-SNE analyses revealed a loss of three additional major triggering receptors required for NK cell activation: NKG2D, DNAM-1, and CD96 (Figure 2A and B). Manual gating confirmed that a significantly lower percentage of CD56^-^CD16^+^ NK cells than CD56^dim^CD16^+^ NK cells expressed the triggering receptors NKp30, NKp46, NKG2D, DNAM-1, and CD96 compared with CD56^dim^CD16^+^ NK cells in patients with AML (NKp30: 14.0 *vs* 47.4%, P<.001; NKp46: 20.5 *vs* 58.9%, P<.05; NKG2D: 36.1 *vs* 84.7%, P<.001; DNAM-1: 35.7 *vs* 71.8%, P<.001; CD96: 21.2 *vs* 42.7%, P<.05) (Figure 2C). No significant variation was observed regarding NKG2C expression (data not shown). Maturation profiles revealed that CD56^-^CD16^+^ NK cells displayed significantly lower expression of maturation markers such as CD57 and Killer cell immunoglobulin-like receptors (KIRs) than conventional CD56^dim^CD16^+^ NK cells; CD57: 27.7 *vs* 61.8%, P<.001; CD158a,h: 9.8 *vs* 19.8%, P<.01; CD158b1,b2,j: 13.8 *vs* 33.7%, P<.001), suggesting that these cells do not represent a senescent subset issued from terminally differentiated NK cells (Figure 3). However, these cells did not belong to an immature subset of NK cells either, since this population poorly expressed NKG2A compared with immature CD56^bright^ NK cells (17.9 *vs* 77.1%, P<.001) or with CD56^dim^CD16^+^ NK cells (17.9 *vs* 47.5%, P<.05) (Figure 3). Most of the variations between CD56^dim^CD16^+^ NK cells and CD56^-^CD16^+^ NK cells were also present in healthy volunteers (Figure 2C and 3). Furthermore, CD56^-^CD16^+^ NK cells displayed reduced expression of the anti-apoptotic proteins BCL-2 and BCL-XL compared with CD56^dim^CD16^+^ NK cells in both healthy volunteers and patients with AML (Figure 4).

**Figure 2.**
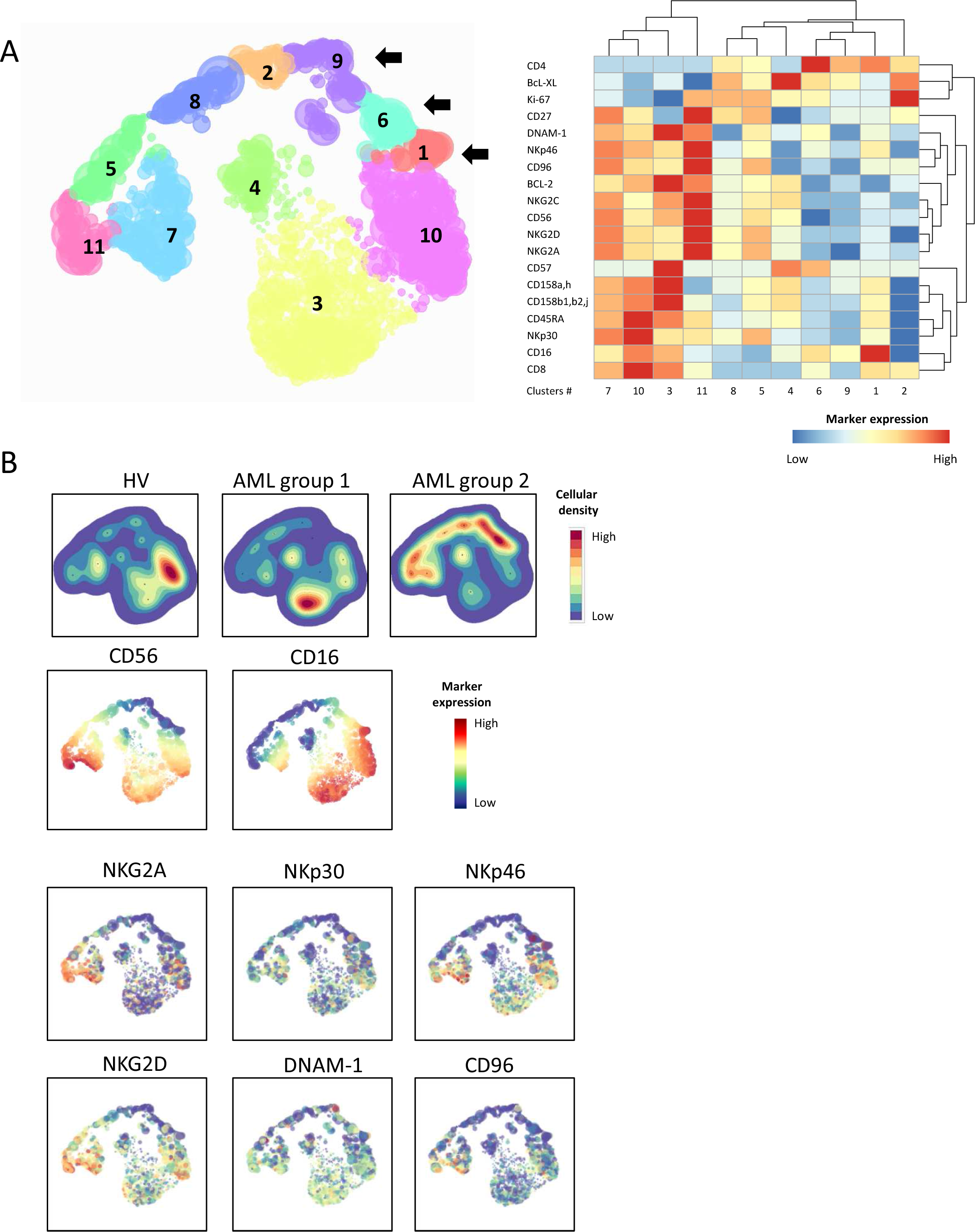
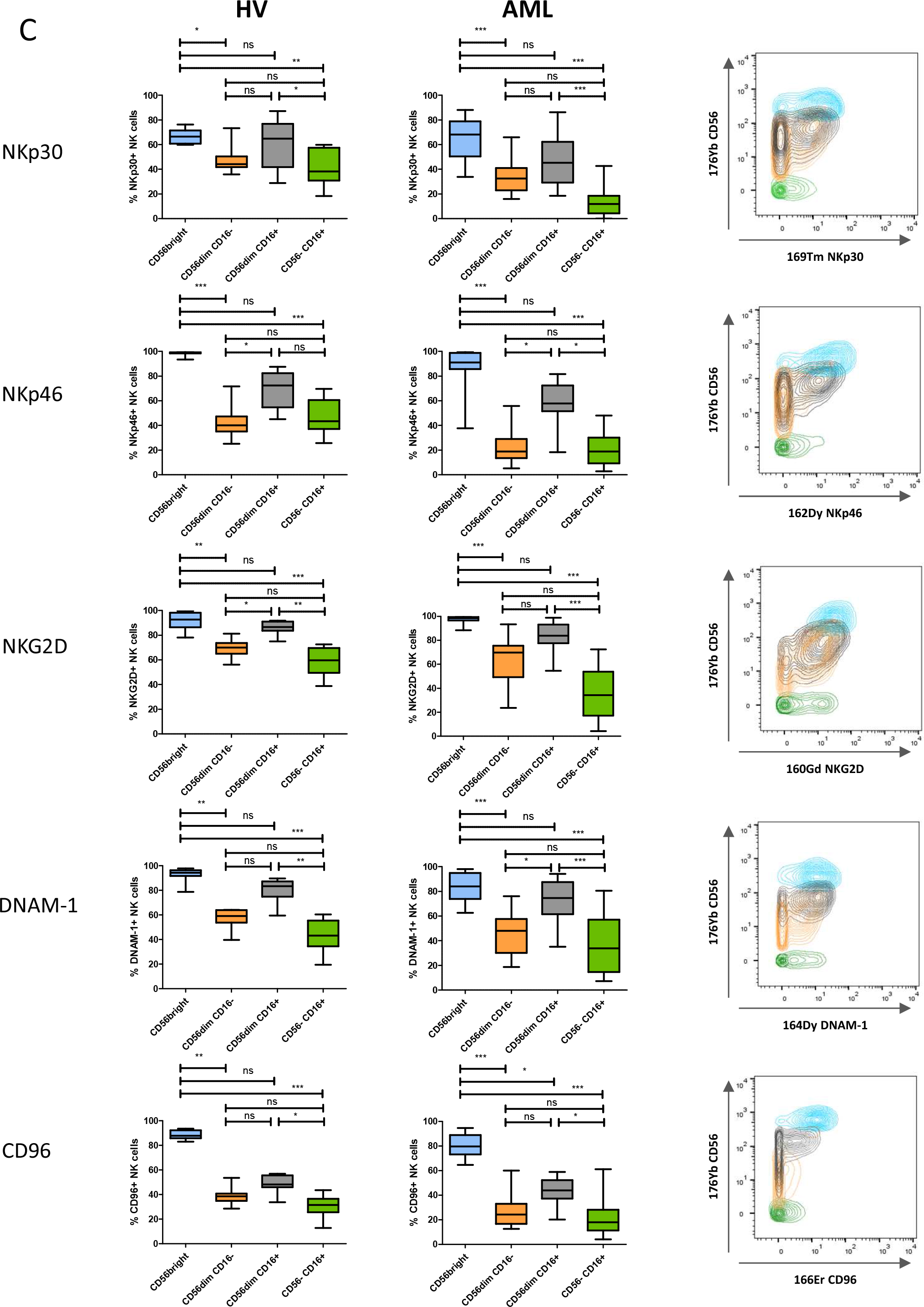
Loss of NK cell-triggering receptors in CD56^-^CD16^+^ NK cells. (A) total NK cells from peripheral blood were manually pre-gated and exported in Cytosplore for h-SNE analysis. Consensus files were generated for each group of patients with fixed number of NK cells. Left panel: h-SNE enables identification of NK cell clusters based on CD56 and CD16 expression; right panel: the heatmap summarizes phenotypical characteristics of NK cell populations identified by h-SNE. (B) Expression of NK cell-triggering receptors projected on h-SNE maps. (C) Left panel: NK cell-triggering receptor expression profiles in clusters of CD56^bright^, CD56^dim^CD16^-^, CD56^dim^CD16^+^, and CD56^-^CD16^+^ NK cells were confirmed by manual gating to enable quantification of differences between clusters of NK cells. Results are presented as interquatile ranges, median, and whiskers from minimum to maximum. Data were analyzed using a Friedman test followed by a Dunn’s test. Right panel: marker expression by NK cell subset; blue: CD56^bright^ NK cells; orange: CD56^dim^CD16^-^ NK cells; grey: CD56^dim^CD16^+^ NK cells; green: CD56^-^CD16^+^ NK cells. *: P<0.05; **: P<0.01; ***: P<0.001; ns: non-significant.

**Figure 3.**
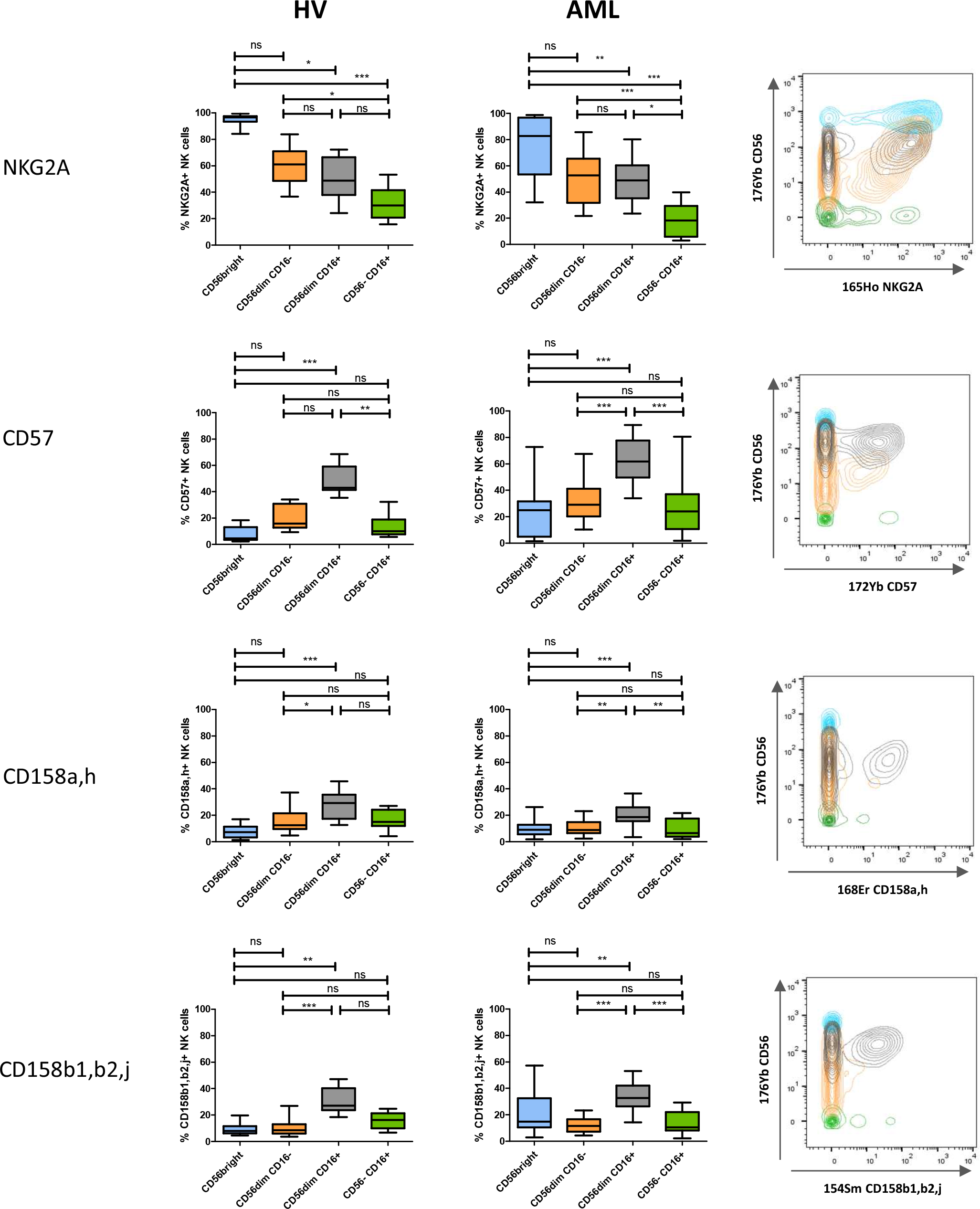
CD56^-^CD16^+^ NK cell clusters display intermediate maturation profiles. Left panel: expression of maturation markers in clusters of CD56^bright^, CD56^dim^CD16^-^, CD56^dim^CD16^+^, and CD56^-^CD16^+^ NK cells was analyzed by manual gating. Results are presented as interquatile ranges, median, and whiskers from minimum to maximum. Differences between clusters were analyzed using a Friedman test followed by a Dunn’s test. Right panel: marker expression by NK cell subset; blue: CD56^bright^ NK cells; orange: CD56^dim^CD16^-^ NK cells; grey: CD56^dim^CD16^+^ NK cells; green: CD56^-^CD16^+^ NK cells. *: P<0.05; **: P<0.01; ***: P<0.001; ns: non-significant.

**Figure 4.**
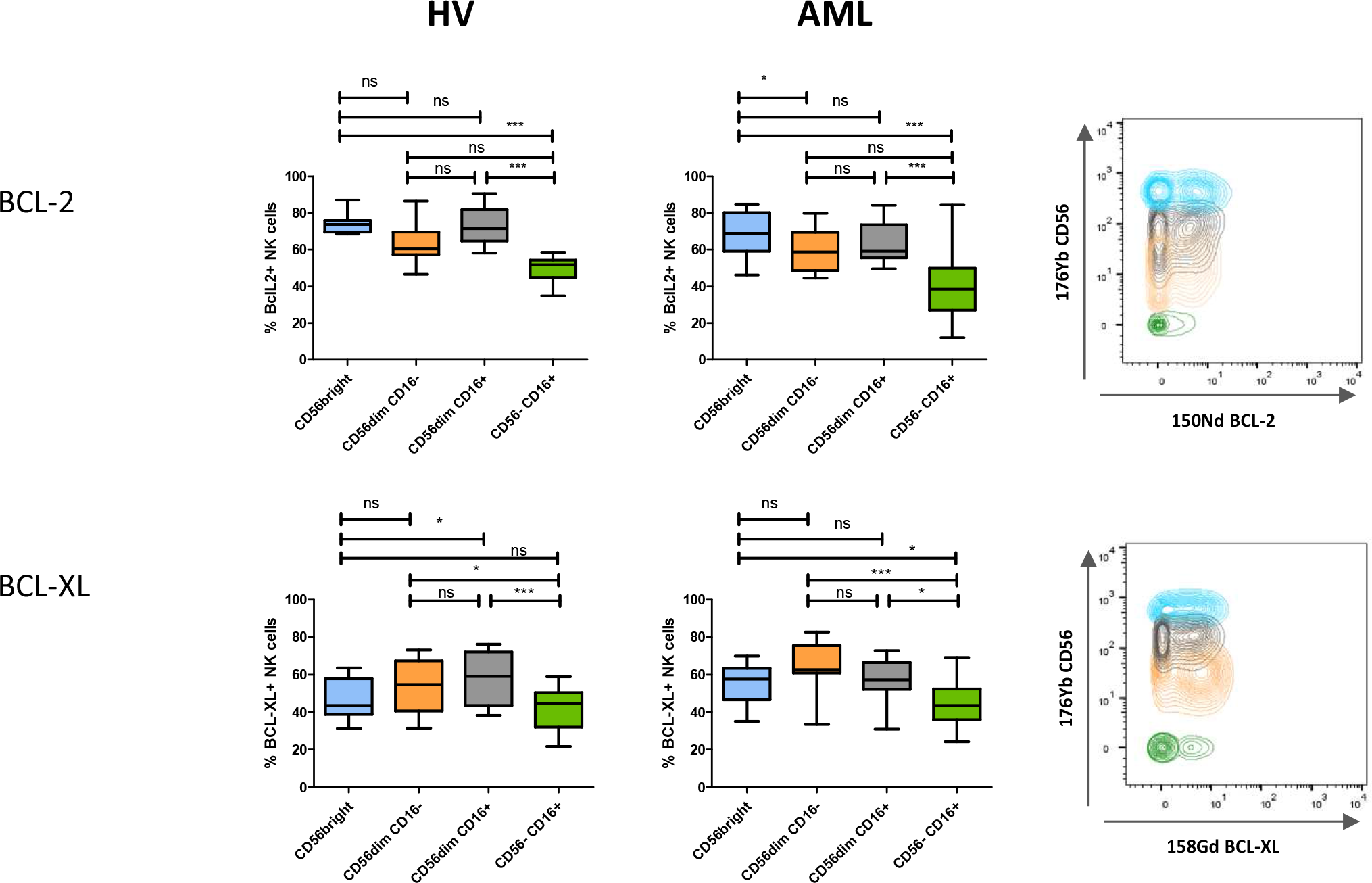
CD56^-^CD16^+^ NK cells display decreased expression of anti-apoptotic proteins. Left panel: expression of anti-apoptotic proteins in clusters of CD56^bright^, CD56^dim^CD16^-^, CD56^dim^CD16^+^, and CD56^-^CD16^+^ NK cells was analyzed by manual gating. Differences between clusters were assessed using a Friedman test followed by a Dunn’s test. Results are presented as interquatile ranges, median, and whiskers from minimum to maximum. Right panel: marker expression by NK cell subset; blue: CD56^bright^ NK cells; orange: CD56^dim^CD16^-^ NK cells; grey: CD56^dim^CD16^+^ NK cells; green: CD56^-^CD16^+^ NK cells. *: P<0.05; ***: P<0.001; ns: non-significant.

### Evolution at complete remission

When material was available, we then assessed variations of the frequency of CD56^-^CD16^+^ NK cells at complete remission compared with baseline frequencies (Supplemental Figure 4). A normal frequency of CD56^-^CD16^+^ NK cells was not systematically restored after induction chemotherapy. After induction chemotherapy, 7 out of 16 patients (43.8%) displayed less than 10% of CD56^-^CD16^+^ NK cells. Among them, 5 patients maintained long-term complete remission and 2 patients relapsed. Nine out of 16 patients (56.2%) displayed more than 10% of CD56^-^CD16^+^ NK cells after induction chemotherapy. Among them, 3 patients maintained long-term complete remission and 6 patients relapsed.

### High frequency of CD56^-^CD16^+^ NK cells at diagnosis is associated with adverse clinical outcome

Patients were classified into two groups according to the frequency of CD56^-^CD16^+^ NK cells, using the thresholds of 10% defined above. The frequency of CD56^-^CD16^+^ NK cells at diagnosis in patients with AML was analyzed according to clinical outcome after 24 months of follow-up. After induction therapy, complete remission rates were 82.9% and 69.2% in groups 1 and 2, respectively. Among patients without accumulation of CD56^-^CD16^+^ NK cells (group 1), 18 out of 35 (54.3%) patients were in continuous CR after 24 months of follow-up, whereas among patients with accumulation of CD56^-^ CD16^+^ NK cells (group 2), 2 out of 13 patients were in continuous CR (15.4%) (Figure 5A). Overall survival (OS) (hazard ratio (HR)=0.13, P<.001) and event-free survival (EFS) (HR=0.33, P<.05) were significantly reduced in patients with high frequency of CD56^-^CD16^+^ NK cells as compared with patients that displayed a conventional NK cell profile, with 3-year OS and EFS rates of 23.7% *vs* 65.9% and 15.4% *vs* 50.4%, respectively (Figure 5B). In multivariate Cox regression models, the frequency of CD56^-^CD16^+^ NK cells retained statistical significance in both OS and EFS (HR=0.19, P=.0004; HR=0.42, P=.033, respectively) (Table 2).

**Figure 5.**
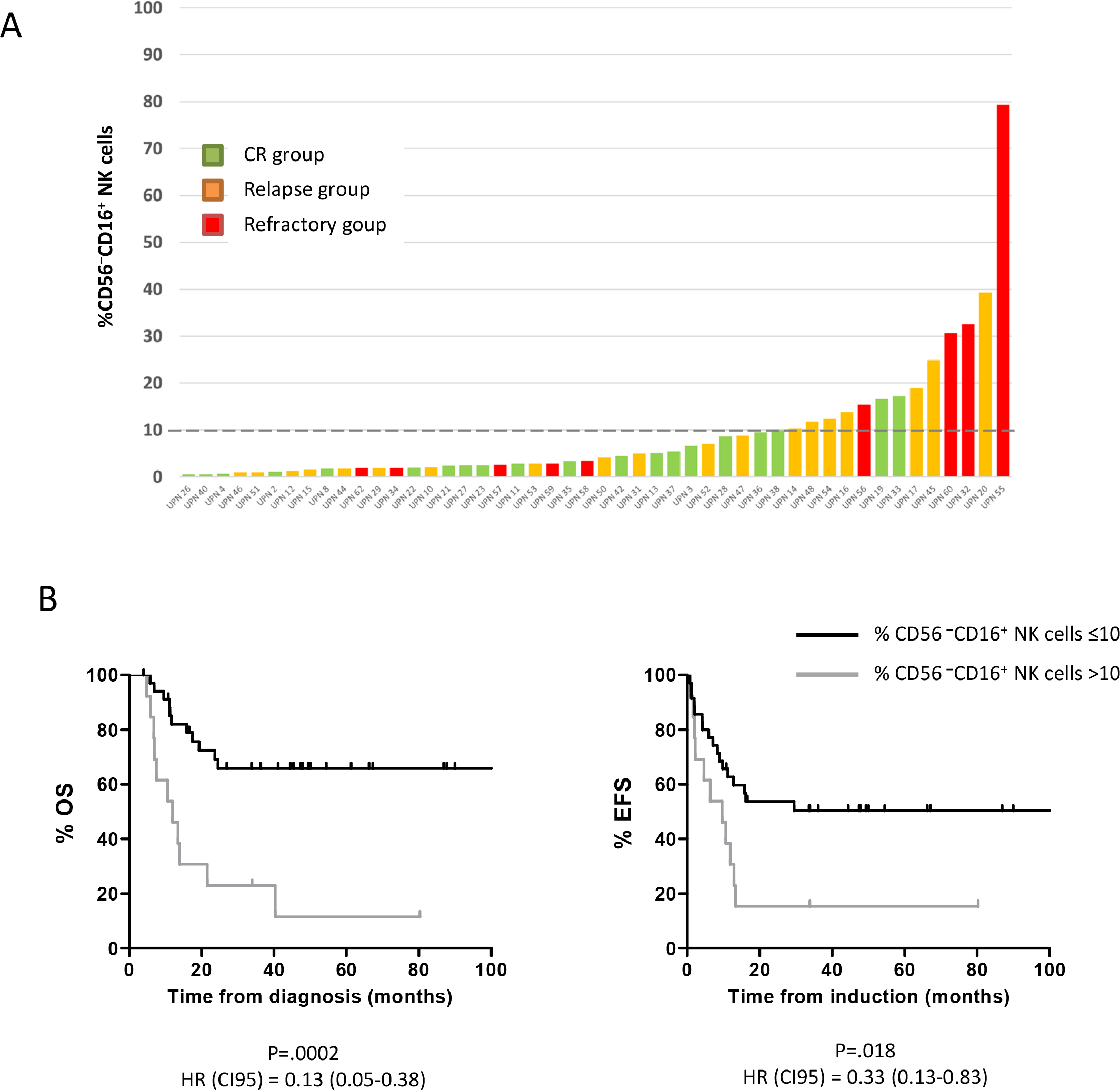
High frequency of CD56^-^CD16^+^ NK cells at diagnosis is associated with adverse clinical outcome. (A) Frequency of CD56^-^CD16^+^ NK cells in AML patients at diagnosis according to clinical outcome after 24 months of follow-up. (B) Patients were stratified according to the frequency of CD56^-^CD16^+^ NK cells (group 1: CD56^-^CD16^+^ NK cells ≤10%; group 2: CD56^-^CD16^+^ NK cells >10%). The impact of the accumulation of CD56^-^CD16^+^ NK cells on OS and EFS was assessed using a Log rank test. CI95, 95% confidence interval; CR, complete remission; HR, hazard ratio; and UPN, unique patient number.

### Pseudo-time analysis

The important question of the origin of these CD56^-^ NK cells remained. Interestingly, in patients with AML, accumulation of CD56^-^CD16^+^ NK cells was associated with a decreased frequency of conventional (CD3^-^CD56^+^) NK cells, thus suggesting that these NK cells are derived from the pool of conventional NK cells rather than expanded from the pool of immature precursor NK cells (Supplemental Figure 5). To probe this hypothesis, we performed a trajectory inference of NK cell maturation using the Wishbone algorithm. Wishbone is a recent algorithm used for analysis of developmental pathways in high-dimensional single-cell datasets. This algorithm positions single cells along bifurcating developmental trajectories on a k-nearest neighbor graph and pinpoints bifurcation points.^32^ The rise and fall of markers acquired or lost during the course of NK cell development (y axis) is represented as a function of pseudo-time (x axis) (Figure 6A and B). Each branch after the bifurcation point represents a distinct differentiation trajectory. Wishbone recovers hallmarks of NK cell maturation in healthy volunteers. NK cells initially highly express CD56 and NKG2A. CD56^bright^ NK cells expressing low levels of CD16 correspond to a transition between early immature CD56^bright^CD16^−^ NK cells and CD56^dim^CD16^+^ NK cells. Subsequently, NK cells lose expression of NKG2A and sequentially acquire KIRs (CD158a,h and CD158b1,b2,j) and finally CD57, which marks the acquisition of high cytotoxic potential^4^ (Figure 6A). In AML, the maturation process is disrupted and leads to a bifurcation point: the first branch displays a normal-like maturation profile, whereas the second branch displays altered maturation features, with decreased CD57 and CD158b1,b2,j expression, as well as loss and re-acquisition of CD16 (Figure 6B). These observations further support the hypothesis of a bifurcation from conventional NK cells toward CD56^-^CD16^+^ NK cells during the maturation process.

**Figure 6.**
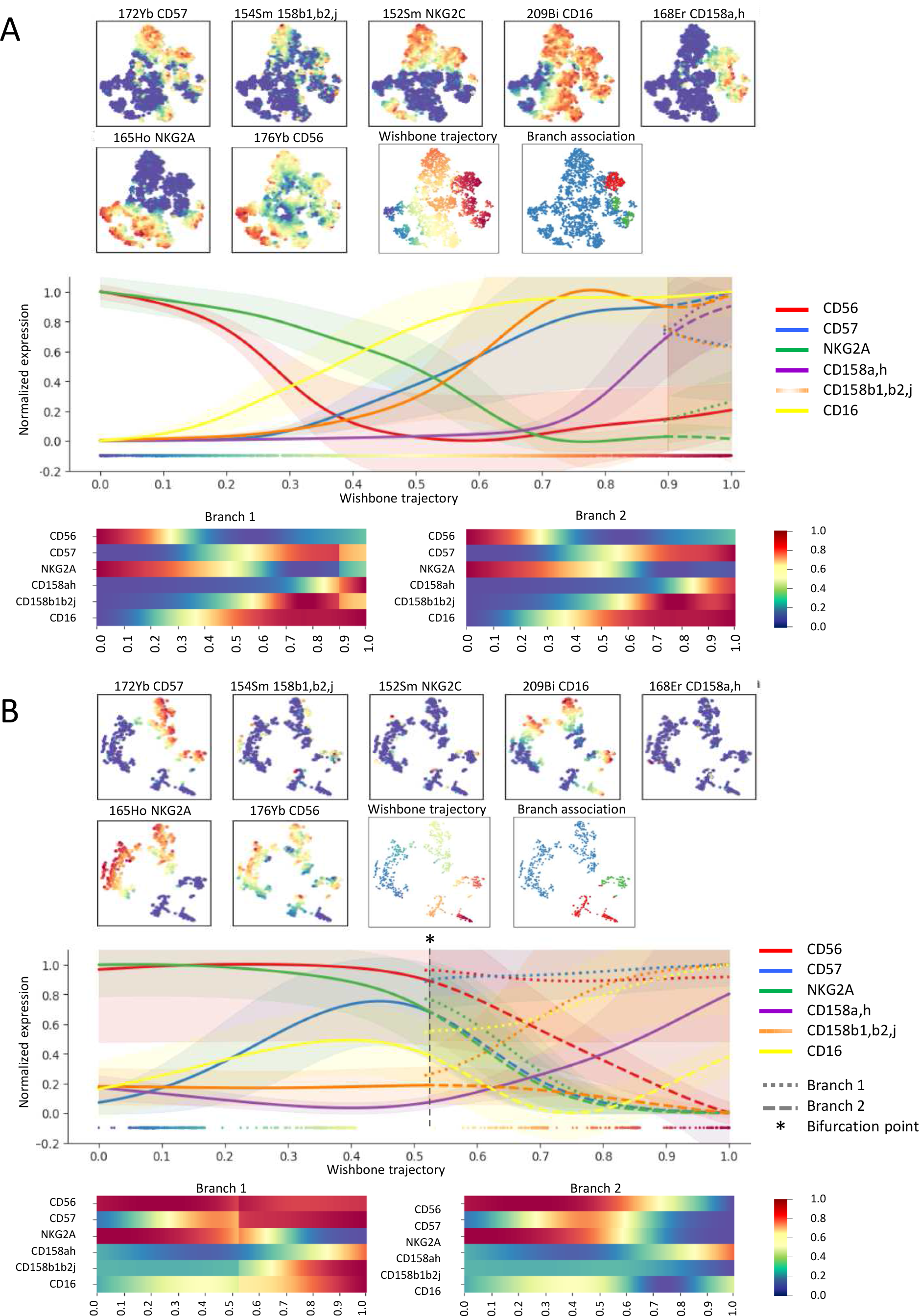
Differentiation trajectories are distinct for CD56^dim^CD16^+^ NK cells and CD56^-^CD16^+^ NK cells. NK cells were manually pre-gated and exported for differentiation trajectory inference using the Wishbone algorithm in HV (A) and AML patients at diagnosis (B). Wishbone enables identification of the branch point that gives rise to the population of CD56^-^CD16^+^ NK cells. Ordering and branching of cells along bifurcating developmental trajectories was performed using t-distributed stochastic neighbor embedding (t-SNE) and diffusion maps. The resulting trajectory and branches are used to visualize the dynamics of maturation markers (CD56, CD57, NKG2A, and KIRs) and of CD16 during NK cell differentiation.

## DISCUSSION

Recent clinical trials highlight the central role of NK cells for the control of AML.^33^ Since NK cell alterations are expected to impact response to NK cell-based immunotherapies, a better characterization of evasion from NK cell surveillance is therefore an absolute prerequisite to design the next-generation of therapeutic strategies based on NK cell manipulation. In the present article, we highlight the presence of unconventional CD56^-^ NK cells in 10% of patients with AML, which is associated with adverse clinical outcomes. This variable appears to be an independent prognostic factor since it retains statistical significance in multivariate analysis. However, allogeneic stem cell transplantation was not equally distributed within groups 1 and 2, with 7 out of 35 patients (20%) *vs* 1 out of 13 (7.7%) receiving allo-SCT, respectively. This difference was not significant and could be partially explained by the lower complete remission rates in group 2.

CD56^-^ NK cells represent a minor subset of NK cells under physiological conditions. Their relative proportion increases in chronically infected HIV viremic patients, in which the CD56^-^CD16^+^ subset can represent up to 50% of total NK cells.^19,20,34,35^ More recently, this population has been described in subjects co-infected with cytomegalovirus (CMV) and Epstein-Barr virus (EBV), with a moderate increase in frequency.^36^ Elevated frequencies of CD56^-^ NK cells have also been described to a lesser extent in other viral chronic infections, such as hepatitis C or hantavirus infections, ^37,38^ and auto-immune ocular myasthenia.^39^ In these pathological situations, functional characterization of these CD56^−^ NK cells revealed profound dysfunctions. In HIV-infected subjects, CD56^-^CD16^+^ NK cells are reported to be highly dysfunctional, with reduced natural cytotoxicity, defective antibody-dependent cellular cytotoxicity, as well as low IFNγ production capacity, although they retain chemokine secretion capacities.^20,21^ Phenotypical characterization of CD56^-^CD16^+^ NK cells highlighted a decreased expression of the NK cell-triggering receptors NKp30 and NKp46, as well as a decreased expression of NKG2A.^20^ Whether this population of NK cells is induced by chronic exposure to antigens is unclear, and mechanisms involved in CD56^-^CD16^+^ NK cell accumulation remain to be elucidated. Overall, previous reports suggest that these NK cells represent a subset of exhausted NK cells.^20,21,35^ Interestingly, an important downregulation of CD56 could be obtained *in vitro* using sera from patients with chronic lymphoid leukemia added to NK cells from healthy subjects, suggesting that this phenotype emerges in the presence of an immunosuppressive milieu.^40^

Beside the question of the mechanisms involved in the emergence of CD56^-^CD16^+^ NK cells, the important question of the origin of this subset remained to be explored. We used a pseudo-time algorithm to explore differentiation trajectories that give rise to CD56^-^CD16^+^ NK cells. These algorithms were developed to investigate the fundamental questions of development processes from individual cells into different cell types. Interestingly, we could reconstitute the theoretical scheme of NK cell maturation in healthy subjects. Identification of a bifurcation occurring during the course of NK cell maturation in patients with AML provides answers regarding the possible origin of CD56^-^ CD16^+^ NK cells, suggesting that accumulation of CD56^-^ NK cells would be the consequence of disruption of the maturation process. This observation provides bases for further exploration of the mechanisms involved in the emergence of this population, which will enable to better define therapeutic strategies likely to restore physiological frequencies of CD56^-^ NK cells. These elements must be considered with regards to other maturation anomalies our group described in AML, with a maturation blockade that affects approximately 10% of patients, for whom the clinical outcome is dramatic.^6^ As with maturation blockade, bifurcation toward CD56^-^ NK cells has major consequences on clinical outcome. Hence, a normal NK cell maturation appears to be critical to control the disease and these anomalies urgently need to be further explored in AML. Interestingly, it has been reported that this phenotype is reversed when the viral load of patients with HIV was lowered below detectable levels after 12 months of antiretroviral therapy, a situation associated with considerable improvement in NK cell function, increased NK cell receptor expression, and restoration of normal CD56 expression.^21^ In the context of AML, the normalization of the frequency of CD56^-^ NK cells was observed only in some patients at the time of complete remission. This persistence of CD56^-^ NK cells 30 days after induction chemotherapy should be explored at later time points to confirm these results.

To our knowledge, accumulation of CD56^-^ NK cells has not been reported in hematological or solid malignancies, except in chronic natural killer cell large granular lymphocytosis (LGL), where there is a massive clonal expansion of CD56^dim/-^ NK cells in a subset of patients.^41^ According to the authors, CD56^-^ NK cell expansion in LGL leukemia might be the consequence of an activating stimulus such as a retroviral infection, notably because sera from patients with LGL leukemia frequently react with HTLV-I/II p21 envelope proteins. Whether CMV infection could induce this aberrant phenotype has not been determined in our cohort, as we do not have CMV status data for all our patients. However, the increase in CD56^-^ NK cells described in CMV and EBV co-infected subjects is moderate^36^ and therefore does not explain the massive accumulation we observe in AML, although a synergistic mechanism cannot be excluded. Finally, we observed a higher prevalence of CD56^-^ NK cell accumulation in AML with inversion 3 (inv 3); in 3 out of 4 patients, CD56^-^CD16^+^ NK cells represented more than 20% of the total NK cell population. This association might represent an interesting feature of immune alteration in the group of patients with inv 3 for whom clinical outcome is extremely unfavorable, and requires confirmation in a larger cohort of patients.^42^

In conclusion, loss of CD56 expression by the NK cells of patients with AML might represent a feature of immune evasion from NK cell control. If the impact of such a phenotype on clinical outcome is confirmed, the frequency of CD56^-^ NK cells at diagnosis might be informative in NK cell-based immune signatures to predict clinical outcome in AML. Lastly, exploration of the mechanisms involved in downregulation of CD56 will provide new opportunities to restore NK cell functions in this specific subgroup of patients.

## Data Availability

The authors provide mass cytometry panels used in the present article as well as source code that were updated for the use of the Wishbone algorithm

https://github.com/moreymat/wishbone/releases/tag/0.4.2-py.3

## ACKNOWLEDGMENTS

The authors thank the CRCM cytometry core facility as well as the IPC/CRCM/UMR 1068 Tumor Bank, which operates under authorization # AC-2007-33 granted by the French Ministry of Research (Ministère de la Recherche et de l’Enseignement Supérieur). Louise Ball from Angloscribe, an independent scientific language editing service, provided drafts and editorial assistance to the authors during preparation of this manuscript. This work has been financially supported by the INCa (grant 2012-064/2019-038 for D.O., and A.T.), the Fondation de France (grant 00076207 for A.S.C), the SIRIC Marseille (grant INCa-DGOS-INSERM 6038), the Cancéropôle PACA (grants K_CyTOF 2014 and AML_CyTOF 2016 for J.A.N.), the GS IBiSA and the Agence Nationale de la Recherche (PHENOMIN project for M.M., H.L., and E.G.). The team “Immunity and Cancer” was labeled “Equipe FRM DEQ 201 40329534” (for D.O.).

## AUTHORSHIP

### Contribution

C.C., C.D., E.G., F.O., M.P., and N.S. performed experiments

A.-S.C., H.L., J.W., M.M., P.R., and S.G., analyzed results and prepared the figures

A.-S.C., A.T., C.F., D.B., D.O., H.L., J.A.N., M.M., N.V., R.D., and T.P. designed the research and wrote the article

