## supplementary data for "High-dimensional mass cytometry analysis of NK cell alterations in Acute Myeloid Leukemia identifies a subgroup with adverse clinical outcome"

**This PDF file includes:**

Figures S1 to S5
Table S1

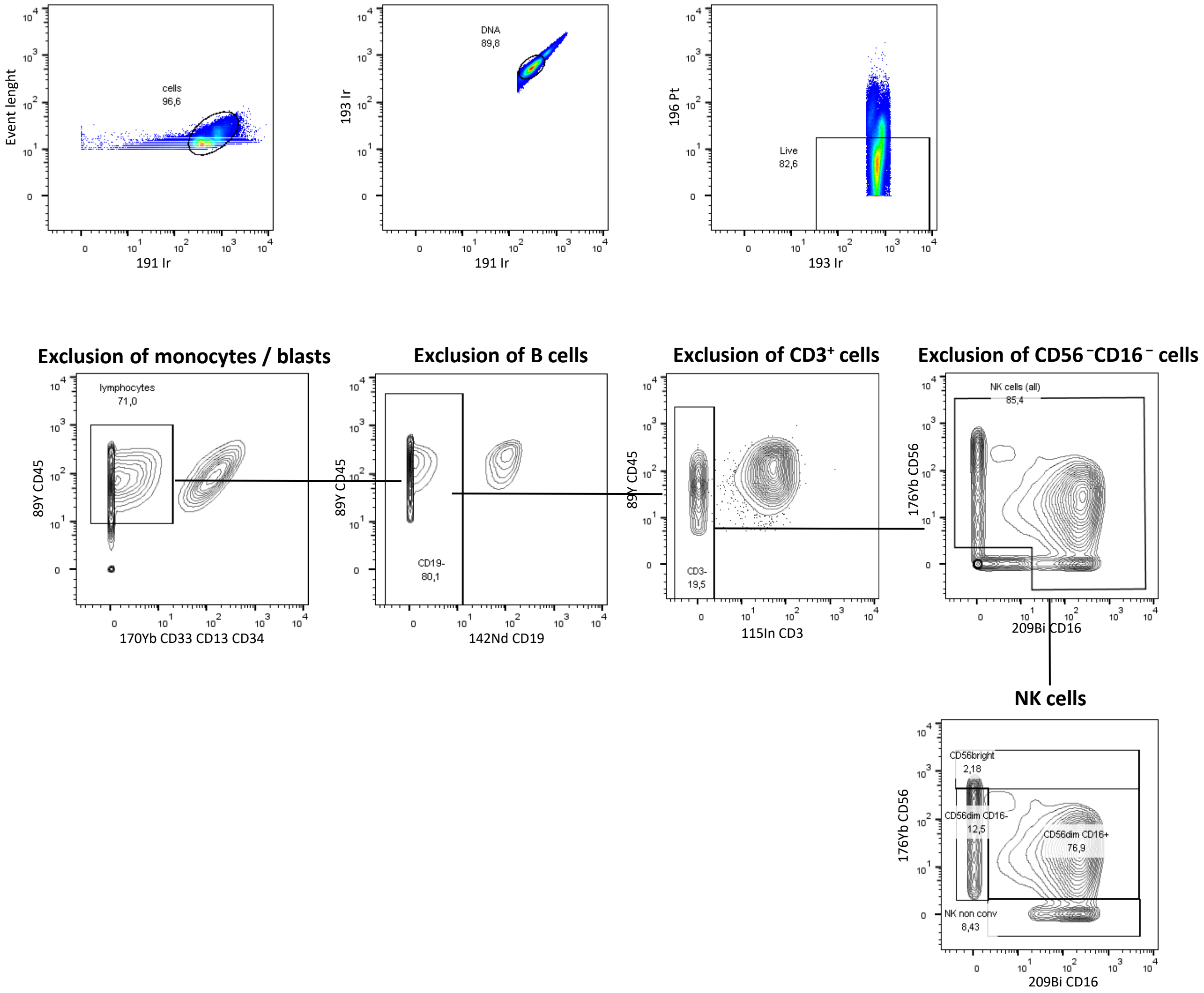


**Supplementary Figure 1. Gating strategy.** PBMCs from HV and patients with AML were stained with the indicated set of antibodies (see Supplementary Table 1) and analyzed by mass cytometry (CyTOF Helios®). After removal of beads, cells were pre-gated as DNA$^{+}$, cisplatin$^{-}$ cells. Monocytes and leukemic blasts were excluded based on expression of CD13, CD33, and CD34 and on low or absent CD45 expression; B cells and T cells were excluded based on CD19 and CD3 expression, respectively. Four subsets of NK cells were defined based on CD56 and CD16 expression: CD56$^{bright}$ NK cells, CD56$^{dim}$CD16$^{+}$ NK cells, CD56$^{dim}$CD16$^{-}$ NK cells, and unconventional CD16$^{-}$CD16$^{+}$ NK cells.

**OS**

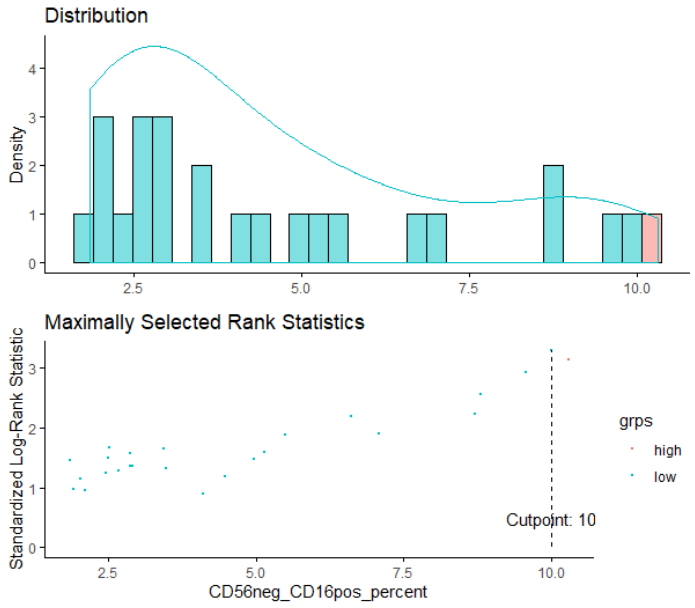


**EFS**

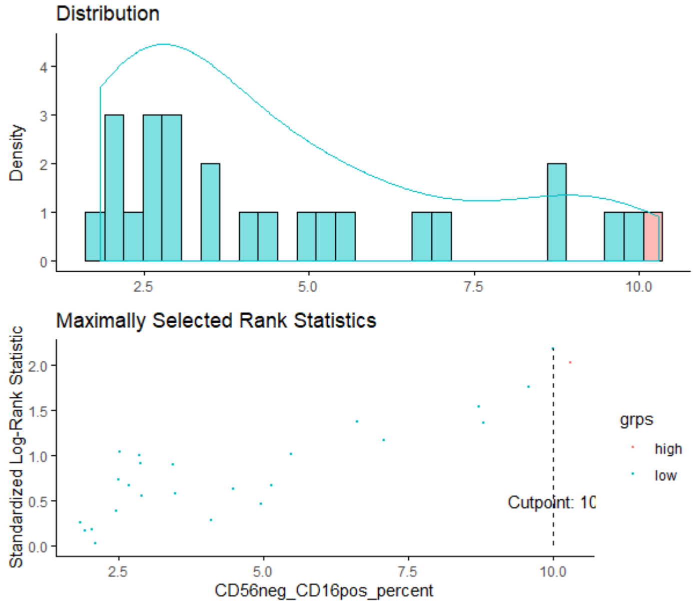


**Supplementary Figure 2. Threshold determination for definition of AML subgroups.** The optimal cut-points were determined using maximally selected log-rank statistics using the maxstat R package. Maxstat enables identification of, for a given prognostic parameter, the optimal threshold that best discriminates two groups of patients.(https://cran.r-project.org/web/packages/maxstat/index.html)

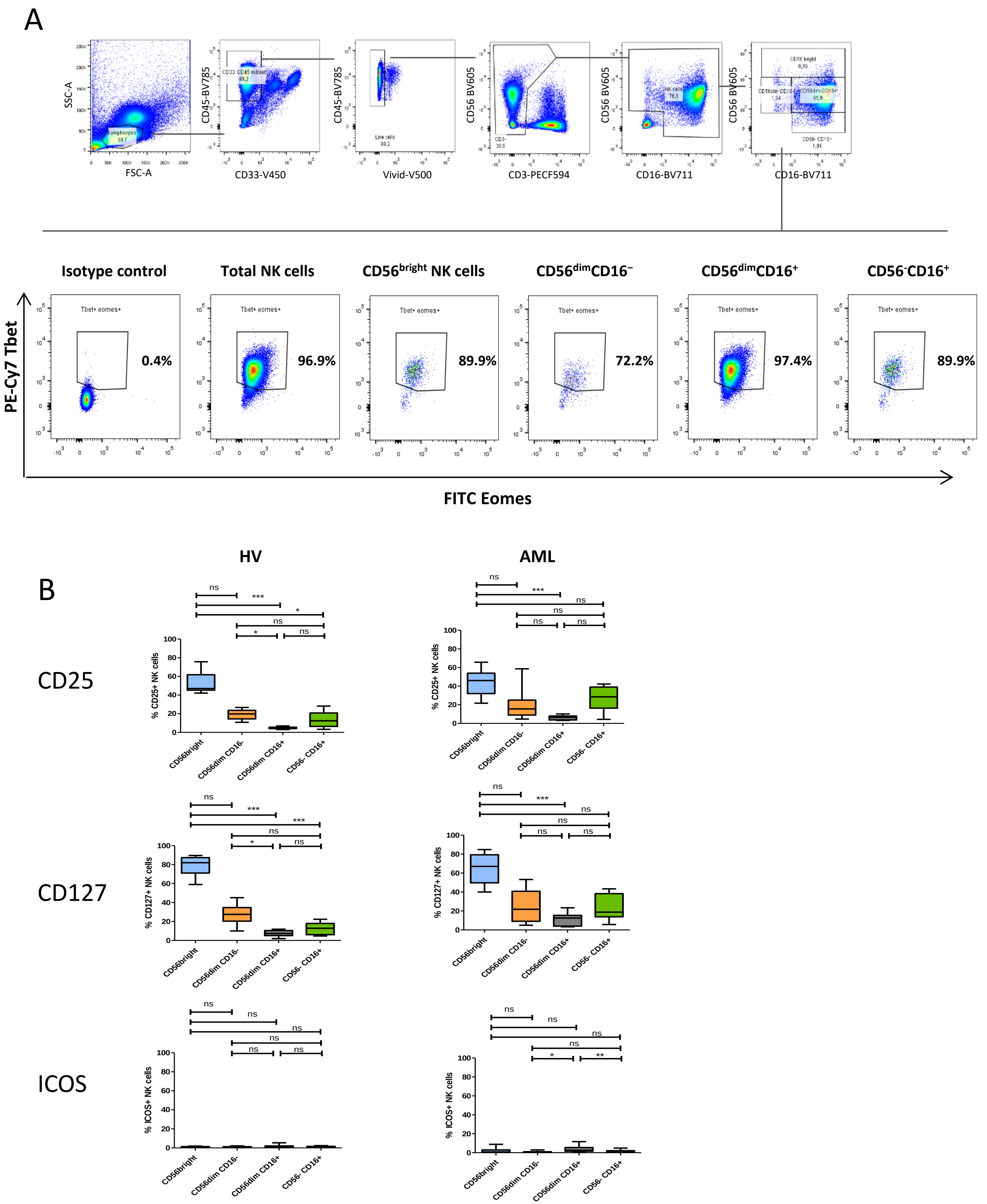


**Supplementary Figure 3. CD3⁻CD56⁻CD16⁺ cells are not group 1 ILCs.** (A) NK cells from HV were analyzed by flow cytometry to assess expression of Tbet and Eomes in the subset of CD56⁻CD16⁺ NK cells. (B) Expression of ILC1 markers (CD25, CD127, and ICOS) was further assessed by mass cytometry in peripheral NK cells from HV and from patients with AML. Differences between clusters were assessed using a Friedman test followed by a Dunn's test.

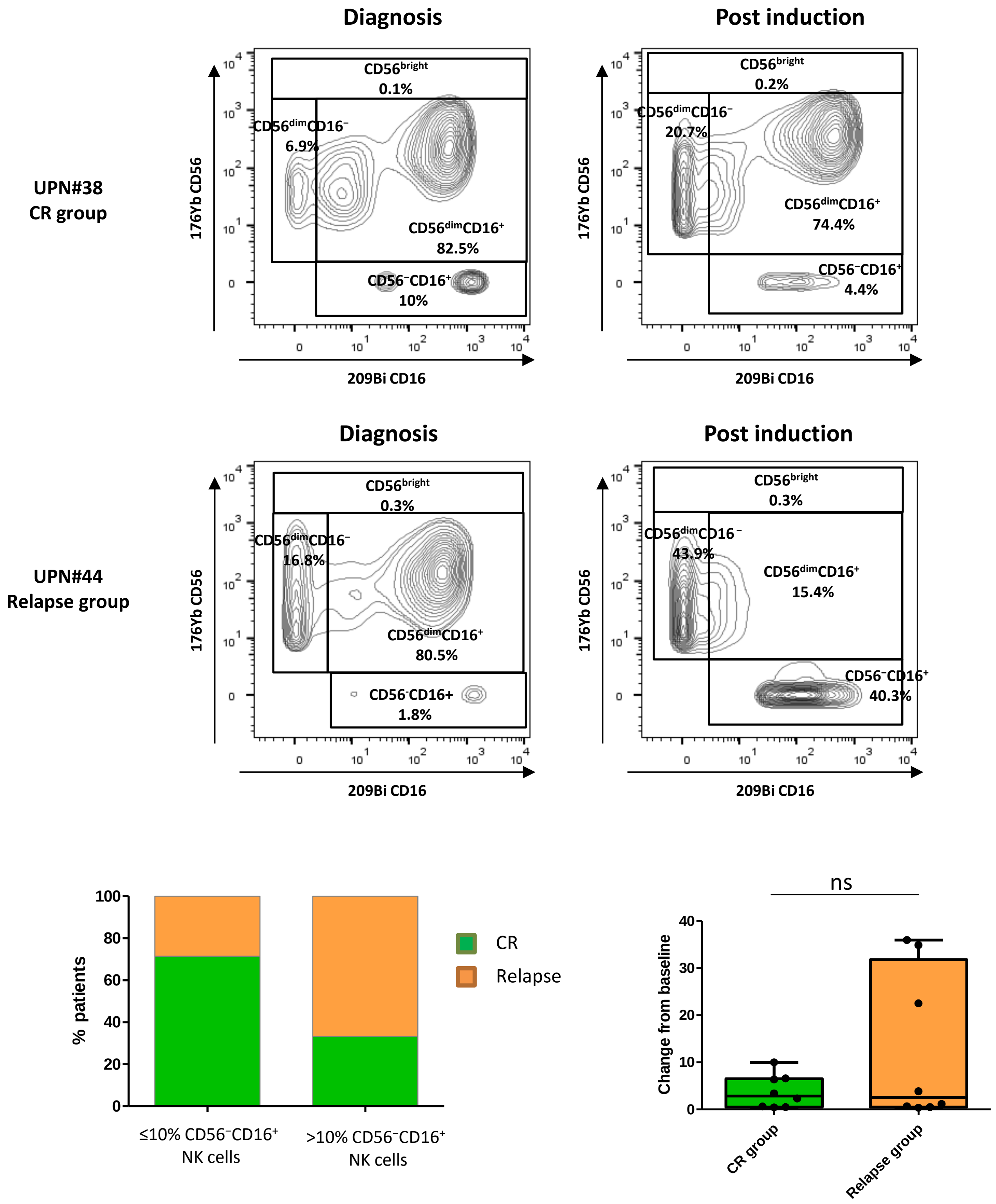


**Supplementary Figure 4. Induction chemotherapy does not restore non-pathological frequency of CD56⁻CD16⁺ NK cells.** The frequency of CD56⁻CD16⁺ NK cells was assessed in paired samples from AML patients at diagnosis and 30 to 45 days after induction therapy (n=16). The change from baseline was calculated as the ratio between the frequency of CD56⁻CD16⁺ NK cells at diagnosis and the frequency of CD56⁻CD16⁺ NK cells at complete remission (CR). Raw frequencies and change from baseline were analyzed by subgroups of patients according to clinical outcome. ns, non-significant; UPN, unique patient number.

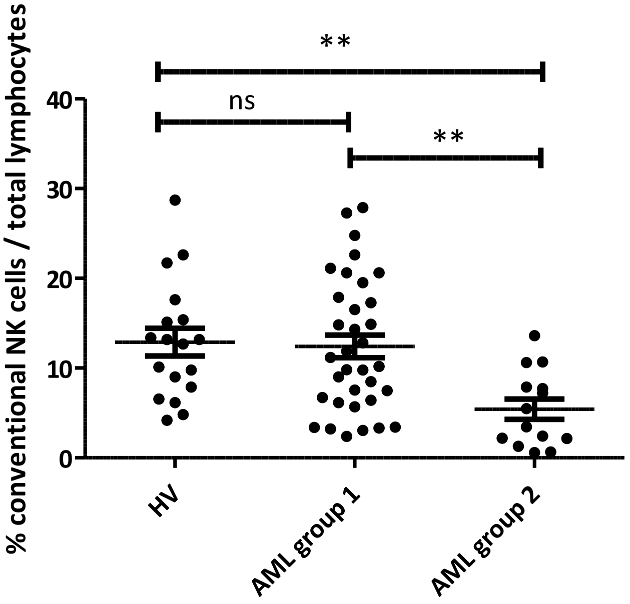


**Supplementary Figure 5. High frequency of CD56$^-$CD16$^+$ NK cells is associated with a decrease of conventional NK cells.** Frequencies of conventional CD3$^-$CD56$^+$ NK cells were determined using manual gating and compared between healthy volunteers, patients with low frequencies of CD56$^-$CD16$^+$ NK cells (group 1) and patients with high frequencies of CD56$^-$CD16$^+$ NK cells (group 2). Results are presented as mean ± SEM. Data were analyzed using a Kruskal–Wallis test followed by a Dunn's post-test. **: P<0.01; ns: non-significant.

### Supplementary Table 1. Mass cytometry panels

Panel 1

| Antigen | Metal/fluorochrome | | Company |
|---|---|---|---|
| **CD45** | 89Y | Extracellular | Fluidigm |
| **CD3** | 115In | Extracellular | CRCM* |
| **TCRVd2** | 141Pr | Extracellular | Beckman Coulter* |
| **CD19** | 142Nd | Extracellular | Fluidigm |
| **CD45RA** | 143Nd | Extracellular | Fluidigm |
| **CD4** | 145Nd | Extracellular | Fluidigm |
| **CD8a** | 146Nd | Extracellular | Fluidigm |
| **BCL-2** | 150Nd | Intracellular | Cell Signaling* |
| **NKG2C** | 152Sm | Extracellular | Miltenyi |
| **TCRpangd** | 153Eu | Extracellular | Miltenyi |
| **CD158b1/b2j** | 154Sm | Extracellular | Beckman Coulter* |
| **CD27** | 155Gd | Extracellular | Fluidigm |
| **BCL-Xl** | 158Gd | Intracellular | Cell Signaling* |
| **Ki-67** | 159Tb | Intracellular | BioLegende |
| **NKG2D** | 160Gd | Extracellular | Beckman Coulter |
| **NKp46** | 162Dy | Extracellular | Fluidigm |
| **DNAM-1** | 164Dy | Extracellular | Beckman Coulter* |
| **NKG2A** | 165Ho | Extracellular | Beckman Coulter* |
| **CD96** | 166Er | Extracellular | BioLegende |
| **CD158a/h** | 168Er | Extracellular | Beckman Coulter* |
| **NKp30** | 169Tm | Extracellular | Beckman Coulter* |
| **CD57** | 172Yb | Extracellular | Fluidigm |
| **CD56** | 176Yb | Extracellular | Fluidigm |
| **CD16** | 209Bi | Extracellular | Fluidigm |
| **CD34** | PE/biotinylated | Extracellular | Beckman Coulter*/Sigma* |
| **CD13** | PE/biotinylated | Extracellular | BD Biosciences*/Sigma* |
| **CD33** | PE/biotinylated | Extracellular | Beckman Coulter*/Sigma* |
| **Anti-PE** | 156Gd | Extracellular | Fluidigm |

Panel 2

| Antigen | Metal/fluorochrome | | Company |
|---|---|---|---|
| **CD45** | 89Y | Extracellular | Fluidigm |
| **CD3** | 115In | Extracellular | CRCM* |
| **Ki-67** | 141Pr | Intracellular | BioLegende |
| **CD45RA** | 143Nd | Extracellular | Fluidigm |
| **HVEM** | 144Nd | Extracellular | Fluidigm |
| **CD8a** | 146Nd | Extracellular | Fluidigm |
| **CD25** | 149Sm | Extracellular | Fluidigm |
| **OX-40** | 150Nd | Extracellular | Fluidigm |
| **ICOS** | 151Eu | Extracellular | Fluidigm |
| **TIM-3** | 153Eu | Extracellular | Fluidigm |
| **TIGIT** | 154Sm | Extracellular | Fluidigm |
| **PD-1** | 155Gd | Extracellular | Fluidigm |
| **PD-L1** | 156Gd | Extracellular | Fluidigm |
| **4-1BB** | 158Gd | Extracellular | Fluidigm |
| **CCR7** | 159Tb | Extracellular | Fluidigm |
| **CD28** | 160Gd | Extracellular | Fluidigm |
| **CTLA-4** | 161Dy | Intracellular | Fluidigm |
| **FOXP3** | 162Dy | Intracellular | Fluidigm |
| **BTLA** | 163Dy | Extracellular | Fluidigm |
| **CD95** | 164Dy | Extracellular | Fluidigm |
| **CD127** | 165Ho | Extracellular | Fluidigm |
| **CD44** | 166Er | Extracellular | Fluidigm |
| **CD27** | 167Er | Extracellular | Fluidigm |
| **CD69** | 168Er | Extracellular | Fluidigm |
| **TCRVd2** | 169Tm | Extracellular | Fluidigm |
| **DNAM-1** | 171Yb | Extracellular | Fluidigm |
| **CD57** | 172Yb | Extracellular | Fluidigm |
| **LAG3** | 175Lu | Extracellular | Fluidigm |
| **CD56** | 176Yb | Extracellular | Fluidigm |
| **CD16** | 209Bi | Extracellular | Fluidigm |
| **CD34** | PE | Extracellular | Beckman Coulter*/Sigma* |
| **CD13** | PE | Extracellular | BD Biosciences*/Sigma* |
| **CD33** | PE | Extracellular | Beckman Coulter*/Sigma* |
| **Anti-PE** | 156Gd | Extracellular | Fluidigm |
